# Early Detection of Autism Spectrum Disorder in Children Using Different Machine Learning Algorithms

**DOI:** 10.1101/2025.04.13.25323013

**Authors:** Sabreen Waheed Kadhum, Mohammed Ali Tawfeeq

## Abstract

Autism spectrum disorder(ASD) is a neurological condition marked by impaired communication abilities, social detachment, and repetitive behaviors in individuals. Global health organization facing difficulties in establishing an effective ASD diagnostic system that facilitates precise analysis and early autism prediction. It is a scientific issue that necessitates resolution. This research presents an approach for the early prediction of children with ASD utilizing significant variables through machine learning (ML) methods. Three stages comprise the suggested technique. First, a 1250-case ASD dataset was identified and preprocessed. Five extremely effective traits with high Pearson correlation coefficient (PCC) are chosen from 10: Sex, Speech delay, Jaundice, Genetic disorders, and family history. Next, chosen ASD feature dataset through its paces using five ML techniques: Naive Bayes (NB), K-Nearest Neighbor (k-NN), Decision Tree (DT), Support Vector Machine (SVM), and AdaBoostM1 (ABM1). The proposed framework is assessed in the third phase utilizing five measurements such as accuracy, precision, predicting time, recall, and F1-score,. The findings revealed that: The NB and K-NN approaches exhibit superior accuracy rates of 99.2% and 97.2%, with minimal prediction times of approximately 0.3 seconds and 0.45 seconds, correspondingly. Conversely, the DT and AdBM1 methods demonstrate a minor decline in accuracy, achieving 94.8% and 87.6%, respectively, along with increased prediction times. Nonetheless, the SVM approach exhibits the least performance, achieving an accuracy of 80.4% with a highest prediction time of 0.84 seconds.

## 1. Introduction

A neurodevelopmental condition, autism spectrum disorder (ASD) impacts around 24.8 million individuals globally. Extremely repetitious interests and behaviors accompany the disorder’s hallmarks of impaired communication and social interaction[1]. Young children with ASD can have considerable functional impairment. Therefore, early and precise diagnosis is crucial for improving quality of life, symptom severity, and maladaptive behaviors. Despite much investigation, ASD neurological processes remain unclear. Therefore, ASD is diagnosed by behavior, not cause or mechanism. After ASD is diagnosed, genetic testing is routinely utilized to uncover hereditary factors, however they merely indicate risk. However, research has advanced significantly[2].

Rigid and repetitious behaviors, poor communication skills, aberrant sensory processing abnormalities, and cognitive inflexibility define the complicated neurological disease known as ASD [3]. Usually showing before the age of three, it influences people all their lifetime. The Center for Disease Control (CDC) estimates that one of fifty-four kids received an ASD diagnosis. Some one percent of the world’s population is thought to have ASD. Early stages of ASD prognosis with great accuracy and high precision are absolutely vital. Children can develop important social and communication abilities with professional care. They can also be eligible for state or federal handicap funds to assist with therapy[4].

The early detection ASD in children within the Iraqi community remains a significant challenge due to limitations in technological infrastructure, inadequate implementation of screening tools, and the general lack of a systemic framework for developmental surveillance. Traditional approaches have proven insufficient to meet the increasing demand for timely diagnosis and intervention, thereby affecting the developmental trajectories of affected children [1]. These constraints are compounded by a general lack of awareness among caregivers and healthcare professionals, limited integration of diagnostic technologies, and the absence of key strategic mechanisms to enhance detection efficacy. Consequently, Iraq continues to experience a high rate of delayed ASD diagnosis, with substantial regional disparities. Empirical evidence indicates that a significant proportion of children in Iraq are not diagnosed with autism until they reach school age or beyond, thereby missing critical windows for early intervention. Moreover, the availability of reliable and standardized national data on autism prevalence, screening rates, and diagnostic outcomes remains limited. According to a 2021 report by the World Health Organization (WHO), children residing in underserved regions lacking specialized pediatric or neurodevelopmental services are at heightened risk of delayed diagnosis and suboptimal care outcomes [4]. From an engineering standpoint, the Iraqi healthcare system faces multiple technological and infrastructural challenges. There is a widespread lack of electronic health records, mobile screening applications, and AI-based developmental assessment tools. Additionally, caregivers and clinicians often lack the technical training required to utilize such systems effectively. Limited financial resources further hinder the acquisition of advanced diagnostic platforms or employment of trained professionals in developmental and behavioral pediatrics. In regions where legacy systems (i.e., paper-based records or non-standardized assessments) are still in use, resistance to integrating modern systems such as telehealth-enabled ASD screening platforms or ML behavior recognition tools persists.

Traditionally, ASD children are predicted by analyzing behavior around 18–36 months. However, delaying intervention at crucial stages of social communication and learning in affected children may reduce its effectiveness. Automated technologies that record a child’s video and voice from public interactions and use machine learning analysis to detect and anticipate high-risk ASD youngsters will make early risk assessment and intervention more accessible. Thus, the new preprocessed and balanced ASD dataset can aid autism research. The preprocessing stages can be considered correct and successfully perform better results than the original ASD dataset [5].

Recent studies underscore the consequences of technological underutilization in early ASD detection. These studies emphasize the necessity for interdisciplinary solutions that merge engineering innovation with public health strategies. For instance, thereby offering data critical for engineering models aimed at scalable, automated ASD predection systems. Following are some of the most innovative approaches for ASD diagnosis:

**Jaber et al.**[6] **2020** Examined the Association Classification (AC) method as a data mining methodology for predicting the presence of autism in individuals. Seven prominent algorithms are chosen to analyze and assess the efficacy of the AC approach regarding its ability to discover correlations among the data. Imbalanced datasets with 21 attributes. **Kaushik et al.**[7] **2021** Utilized ML algorithms including Support Vector Machines (SVM), Random Forest Classifier (RFC), Naïve Bayes (NB), Logistic Regression (LR), and K-Nearest Neighbors (KNN) to develop prediction models based on the results. The principal constraint of this research is the limited accessibility of extensive and open-source ASD datasets. A substantial dataset is essential for constructing an accurate model. The dataset utilized herein lacked an enough number of cases. **Uzma et al.** [8] **2021** proposed an autistic gesture identification system utilizing wearable biosensors and classification techniques. The proposed system delineates, observes, and categorizes distinct gestures. Utilizing Bluetooth-enabled wearable biosensors to transmit data to a data collection and classification server. Each movement is executed 10 times by 10 autistic adolescents to create a dataset of 24 gestures. Features in the time and frequency domains derived from sensor data are classified utilizing KNN, DT, NN, and RF algorithms. The primary limitation of this study is that children with ASD executed movements while in standing positions, which is restrictive. The technology has not undergone testing with biosensors from children with ASD in various body positions. A solitary wrist biosensor was employed to collect data, hence intricate gestures cannot be identified. **Khairan et al.** [9] **2021** used attribute selection techniques, chose three classification algorithms— AdaBoost, kNN, and ID3—to build models utilizing datasets chosen ahead to training. Furthermore, show which attribute selection methodologies find pertinent characteristics influencing the first diagnosis process. These approaches directly influence the categorization of screening tools. ML models were used to assess the efficiency of features chosen from an actual toddler dataset using CFS, IG, GI, CHI, and FCBF. **Amandeep and Karanjeet** [10] **2022** Employed six distinct ML algorithms for classification, including C4.5, k-Nearest Neighbors, Random Forest, Logit Boost, Support Vector Machine, and Naive Bayes, to precisely identify ADHD in adults utilizing real-time activity data. The primary limitation is the paucity of data samples resulting from challenges in clinical data gathering. **Mousumi et al.** [11] **2022** Applied several classifiers into datasets of toddlers, children, teenagers, and adults. Besides, used several feature selection methods to support these baselines and more effectively feature sets. This model studied important features but was not trained using different multivariate/dimensional datasets. **Anita and Dipti** [12] **2023** built a multi-classification model that included multiple classifiers in order to improve the accuracy of the autism spectrum disorder prediction. The performance of the model was evaluated using a number of different machine learning techniques, which were then applied to three datasets consisting of children, adults, and adolescents that were not balanced. **Dhuha and Saja** [13] **2023** Utilized various predictive algorithms on four accessible non-clinical ASD screening datasets. These datasets encompass children, adolescents, toddlers, and adults. Models for ASD identification was assessed based on various performance metrics. Examples include DT,K-NN, DT, NBs, and RF. **Muhammad et al.** [14] **2023** utilized Federated learning (FL) approach for ASD detection by training with two distinct ML-based classifiers: SVM and LR to classify ASD variables and detection children as well as adults with ASD. Furthermore, challenging to apply the proposed model for more complicated tasks requiring deep neural networks or other advanced ML models is the complexity of the proposed architecture in addition to training federated learning models on several devices with limited processing capability and storage. Moreover, difficult to create models that operate well across all devices is the heterogeneity of the suggested data dispersed among several devices and locations whereby different devices have different kinds of data. **Anupam et al.** [15] **2025** leveraged a varied array of foundational models, including LR, K-NN, NNs, DT, and SVM for feature selection. A crucial component of the methodology is the integration of complex feature selection and model stacking techniques. The emphasis is on determining the four primary characteristics most diagnostic of ASD for future training in early detection of ASD in toddlers.

ASD diagnosis and prediction research have shown many substantial limitations and challenges. One issue is that small homogeneous sample sizes reduce the generalizability of results to other populations. Subjective assessments like clinical observations and parent reports are used in many studies, which can skew results. Since certain models lack interpretability and durability, limited use of advanced machine learning approaches is another issue. The focus on distinct features rather than biological, environmental, and behavioral factors has also led to limited comprehension. These issues must be addressed to improve ASD diagnosis and prediction. ASD diagnosis requires clinical judgment and standardized tests to ensure precise diagnosis and appropriate treatment. In child healthcare, autism is a major obstacle. ASD identification is crucial for early interventions that may impact development.

This paper utilizes medical records of 1250 cases with ten different features, such as (age, sex, Speech delay, learning disorder, genetic disorders, depression, social issues, anxiety disorder, jaundice, and family history). The proposed algorithms predict children’s cases into two cases, ASD and non-ASD. The prediction of these cases is based on the selected data using five ML algorithms: NB, k-NN, DT, SVM, and AdaBoostM1 (ABM1) constructed predictive methods based on group of ten features representing the longitudinal reversal of ASD that learns from labeled instances to categorize new, unseen cases. A diversity of features offers valuable information, potentially enhancing model accuracy. If the features represent significant elements of the data, the model will more effectively identify patterns. Conversely, if the features are extraneous or contain noise, they may result in diminished accuracy. This arises from a phenomenon known as "overfitting," wherein the model acquires non-beneficial patterns from the training data, hence impairing its performance on novel data. Consequently, employing a strategy like dimensionality reduction is crucial for identifying the most advantageous features and improving model performance.

There are a number of ways to determine the correlation between attributes in a dataset to comprehend the interdependencies and linkages among them within the framework of feature analysis for ML. Enhance model performance by combining features according to their correlations. Among the most effective methods is Pearson Correlation Coefficient (PCC) which is measures the linear relationship between two continuous variables. The PCC is widely used in statistics, research, and data analysis to understand relationships between variables[16]. Provides the ability to remove redundant features (features are highly correlated), consider removing one to reduce multicollinearity as well as select relevant features by choose features that have strong correlation with the target variable (dependent variable) and lower correlation with each other.

This study not only facilitates the identification of personalized delays in brain function or structure linked to ASD but also illustrates the efficacy of pertinent data in elucidating the relationships among features regarding their impact on model performance for predictive models of early ASD indicators in physiological data samples, which has been the emphasis of ASD prediction efforts via the utilizing of PCC method with diverse attributes to enhance precision measurements.

The five sections of this work are: Section 2 shows the suggested technique in great depth and introduces the adopted prediction models to be used. Section 3 addresses the evaluation standards. Section 4 presents the results drawn along with compression to state of art studies. Section 5 clarifies last remarks derived from the main results of the work and possible future paths.

## 2. Methodology (System Model)

The main objectives of this research are to predict the probability of ASD at an early stage and to further delineate the attributes that contribute more significantly than others to the prediction of enhanced precision. This may reduce the costs associated with several patient trials, as not all characteristics may significantly influence the outcome prediction.

Based on the dataset, the suggested framework for the early prediction of ASD among clinical controls is presented in Figure 1 and is therefore helpful for early prediction of whether a child will or not acquire ASD. Data is first obtained from the Kaggle site [17], which comprises patient information including background of the "sociodemographic and family history " risk factors.

**Fig 1:**
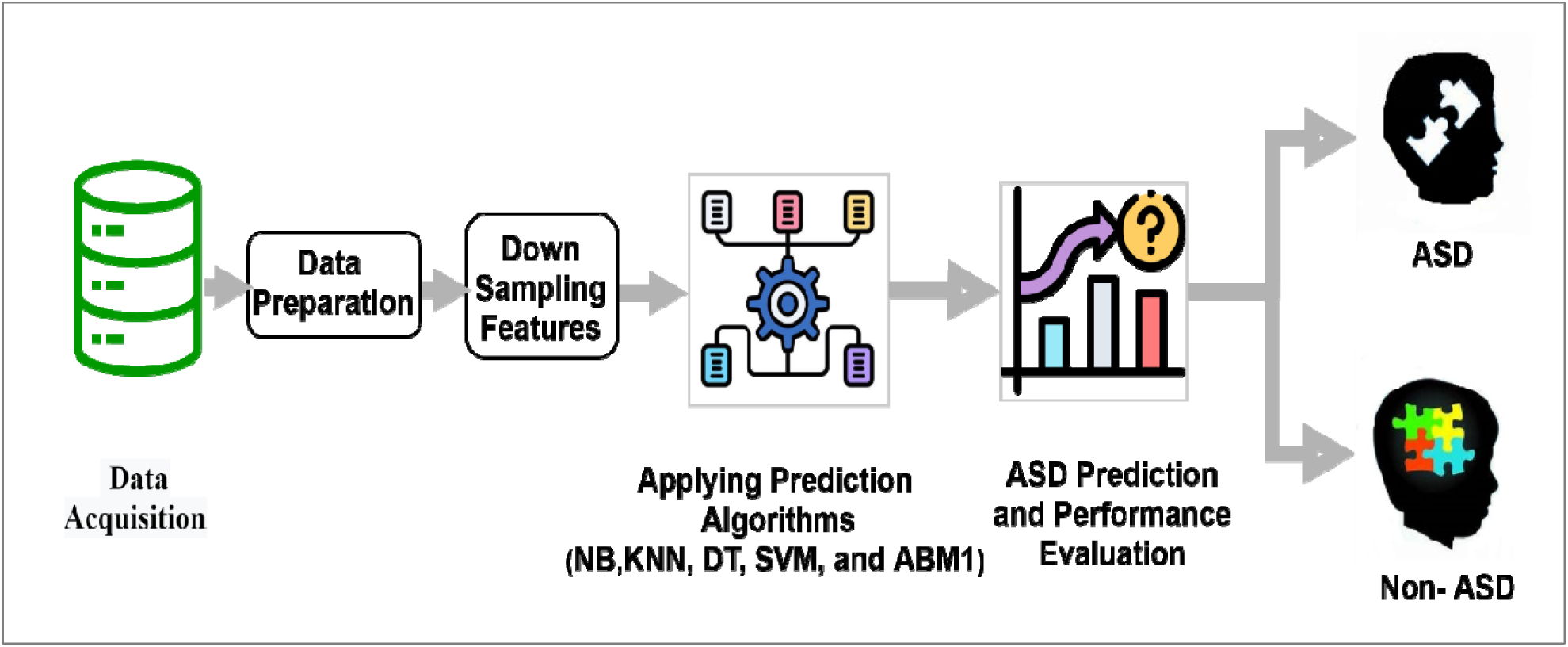
Proposed framework

PCC technique was implemented to find the correlation among ten features. Five distinct features are extracted with highest positive correlation value and then fed into five different ML algorithms (NB, k-NN, DT, SVM, and ABM1) for farther process.

### 2.1 Data Acquisition and Preprocessing

During the data partitioning procedure, the dataset including 1250 instances is segmented into two segments for further analysis. 80% of the data is designated for training, while the remaining 20% is reserved for testing. The data in the first column of Table 1, pertaining to gender, is encoded as zero for female and one for male. The phrases "yes" and "no" are assigned the values one and zero, respectively, throughout the data collection.

**Table 1:** Samples of the implemented dataset.

| Case | Sex | Speech Delay | Learning disorder | intellectual disability | Social Issues | Depression | Anxiety disorder | Jaundice | Genetic disorder | Famil y_hist ory | ASD_ traits |
| --- | --- | --- | --- | --- | --- | --- | --- | --- | --- | --- | --- |
| 1 | 0 | 1 | 1 | 1 | 1 | 1 | 1 | 1 | 1 | 0 | 1 |
| 2 | 1 | 1 | 0 | 1 | 0 | 1 | 1 | 1 | 1 | 1 | 1 |
| 3 | 1 | 1 | 1 | 1 | 1 | 1 | 1 | 1 | 1 | 0 | 1 |
| 4 | 1 | 1 | 1 | 1 | 1 | 1 | 1 | 0 | 0 | 0 | 1 |
| 5 | 0 | 1 | 1 | 1 | 1 | 1 | 1 | 0 | 0 | 0 | 0 |
| 6 | 1 | 1 | 1 | 1 | 1 | 1 | 1 | 0 | 0 | 0 | 1 |
| 7 | 1 | 1 | 1 | 1 | 1 | 1 | 1 | 1 | 1 | 0 | 1 |
| 8 | 1 | 1 | 1 | 1 | 1 | 1 | 1 | 1 | 1 | 0 | 1 |
| 9 | 1 | 1 | 1 | 1 | 1 | 1 | 1 | 0 | 0 | 0 | 0 |
| 10 | 1 | 1 | 1 | 1 | 1 | 1 | 1 | 0 | 0 | 0 | 1 |
| 11 | 1 | 1 | 1 | 1 | 1 | 1 | 1 | 1 | 1 | 1 | 1 |
| 12 | 1 | 1 | 1 | 1 | 1 | 1 | 1 | 1 | 1 | 0 | 1 |
| 13 | 0 | 1 | 1 | 1 | 1 | 1 | 1 | 1 | 1 | 0 | 0 |
| 14 | 0 | 1 | 1 | 1 | 1 | 1 | 1 | 1 | 1 | 0 | 1 |
| 15 | 1 | 1 | 1 | 1 | 1 | 1 | 1 | 1 | 0 | 0 | 0 |
| 16 | 1 | 1 | 1 | 1 | 1 | 1 | 1 | 1 | 0 | 0 | 1 |
| 17 | 1 | 1 | 1 | 1 | 1 | 1 | 1 | 1 | 1 | 1 | 0 |
| 18 | 0 | 1 | 1 | 1 | 1 | 1 | 1 | 1 | 1 | 0 | 1 |

### 2.2. Feature Selection

A crucial phase in the evolution of prediction systems, feature reduction improves performance and efficiency alike. Feature reduction reduces the dimensionality of the dataset by removing pointless or duplicate elements, therefore enabling shorter training and inference times. Moreover, feature reduction can help to increase model interpretability, thereby facilitating the understanding of the feature-predicted connection. Techniques like PCC are frequently utilized to evaluate the correlation among attributes while maintaining the dataset’s integrity. Ultimately, efficient feature reduction allows ML algorithms to concentrate on the most pertinent information. In this research, PCC approach is involved to likelihood among the ten features (sex, speech delay, learning disorder, intellectual disability, social issues, depression, anxiety disorder, jaundice, genetic disorder, and family history).

The PCC, often denoted as (r), measures the strength and direction of the linear relationship between two continuous variables. This value ranges from [-1 to +1]: here’s how it works[18],[19]

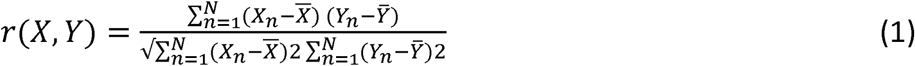

Where:

N is the number of samples, X and Y are two features being compared, and X̄ and Ȳ represents the mean of specific features respectively.

By compute the Pearson correlation coefficient (r) between each pair of features in dataset.

r = 1:Perfect positive correlation

r = -1:Perfect negative correlation

r = 0:No correlation

Construct a correlation matrix displaying the feature pairs’ correlation coefficients. Strong correlations can be better seen and identified with the aid of this matrix. Table 2 displays the association among the 10 features that were implemented.

**Table 2:**
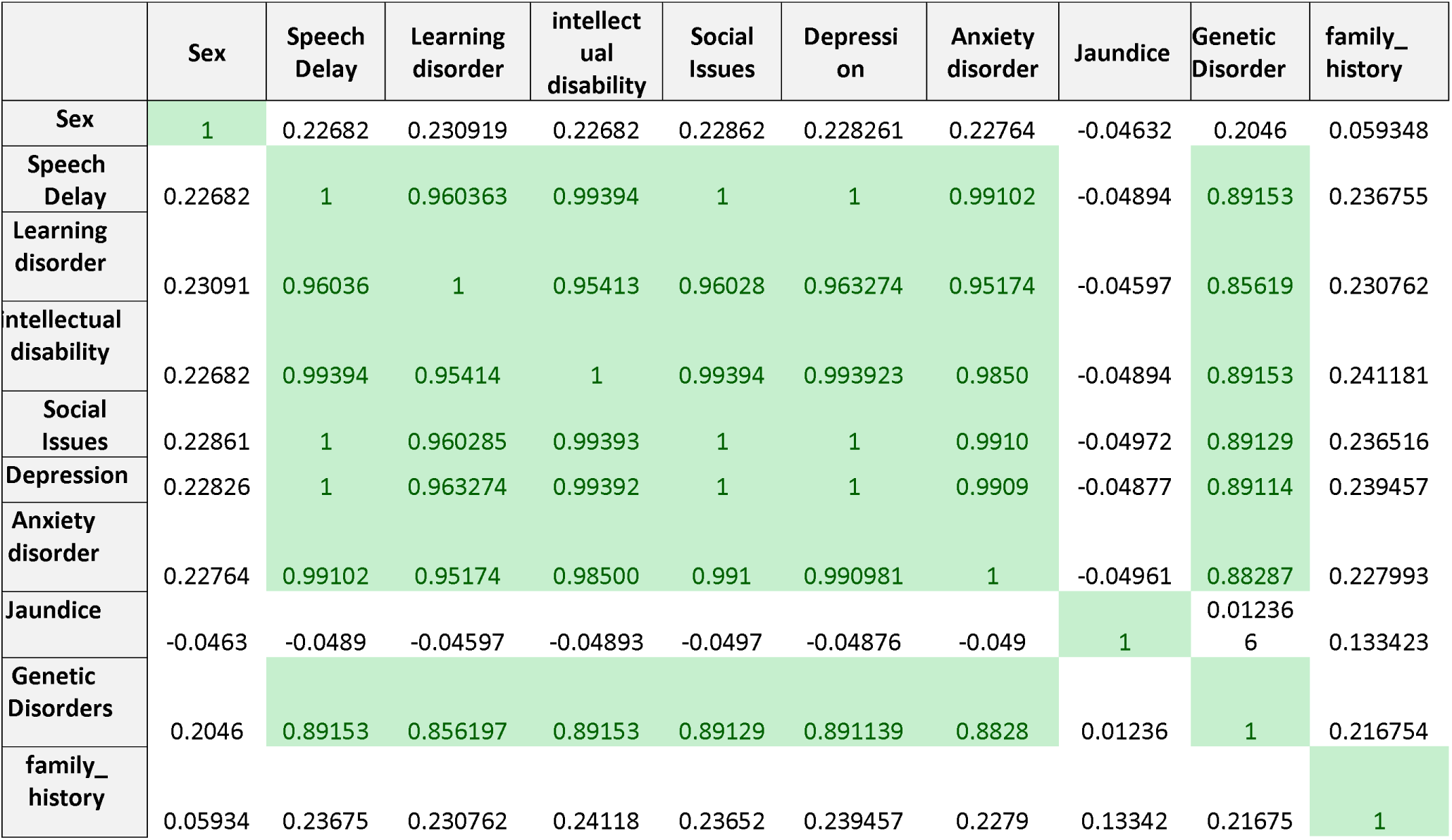
Correlation matrix for a dataset with ten features.

There is a notable positive correlation value between (**speech delay, learning disorder, intellectual disability, social issues, depression,** and **anxiety disorder**) around (0.981). whereas, ‘**Jaundice**’ has the highest negative correlation around by (− 0.05) indicating an inverse relationship with all nine features. Understanding these connections is crucial for genetic counseling, risk assessment, and early intervention in families with a history of ASD or related conditions.

To summarize, selecting the appropriate correlation approach is determined by the types of datasets used and the distribution assumptions used. Each technique has its own merits and is appropriate for various sorts of analysis.

### 2.3 Prediction

This study used multiple supervised ML methods. The fundamental algorithms are largely trained using the labeled dataset. The qualifying model categorizes a non-labeled testing dataset into appropriate categories. The next subsections provide recommended supervised ML techniques for disease detection[13] [10]

#### 2.3.1 Naive Bayes (NB)

NB is a straightforward and effective classification technique grounded in Bayes’ theorem, presupposing the independence of features. The method computes the probability of each class based on the characteristics and designates the label with the greatest likelihood. Although it performs exceptionally well with small datasets and has rapid computing speed, its efficacy may diminish when characteristics exhibit significant correlation. Naïve Bayes continues to be a favored option due to its simplicity and efficacy.

#### 2.3.2 K-nearest neighbor(k-NN)

K-NN is one of the simplest classification algorithms. K is the number of nearest neighbors, which can be supplied in the object constructor or calculated using the value’s upper limit [24]. Thus, analogous instances are classed similarly, and a new case is classified by comparing it to each. After receiving an unidentified sample, the closest neighbor algorithm will search the pattern space for the k training samples next to it. The distance from the test instance may be used to compute predictions from numerous neighbors, and two methods are shown to weight the distance.

#### 2.3.3 Decision Tree (DT)

DT is a versatile and intuitive ML algorithm used for classification and regression tasks. It works by splitting data into subsets based on feature values, creating a tree-like structure of decisions and outcomes. Each intermediate node represents a test on a feature, branches correspond to possible outcomes, and leaf nodes provide the final prediction. DT are easy to interpret and handle both numerical and categorical data effectively. The DT model makes analysis based on three nodes.

- Base node: main node, based on this all-other nodes functions.
- Intermediate node: handles various attributes.
- Leaf node: represent the result of each test.

#### 2.3.4 Support Vector Machine (SVM)

A supervised ML approach, SVM are utilized for prediction, regression, and classification applications. possesses a high degree of efficiency in high-dimensional spaces and is resistant to overfitting, particularly in situations when the number of features is greater than the number of samples. As a result of the decision function using just a portion of the training data, SVM are efficient.

#### 2.3.5 AdaboostM1 (ABM1)

Ensemble learning-based supervised ML classifier ABM1. It uses adaptive improvement to combine many weak classifiers into a robust classifier for better classification results. The introductory phase weights all observations equally. Weak classifier coefficients affect observation weights, whereas estimate error values determine classifier coefficients. Classifier coefficients are their error values. Thus, the ABM1 algorithm can weight misclassified data higher and well-known ones lower. In subsequent rounds, it will emphasize misclassified observations. By employing linear combination, all weak classifiers are combined to build a more robust classifier that produces accurate classification results. The ASD dataset is trained and evaluated using these approaches. The Kaggle library runs prediction algorithms.

## 3. Evaluation Metrics and Performance Analysis

Five predictive algorithms were employed on an analogous balanced dataset including 1250 instances to identify the most effective method by comparing accuracy and other statistical metrics. The algorithms utilized were(NB, KNN, DT, SVM and ABM1). This article provides a concise overview of the performance reviews. Table 3 illustrates the confusion matrix of the proposed study to assess the performance of each deployed method.

**Table 3.** Confusion matrix for ASD prediction.

|  | Child with ASD | Child without ASD |
| --- | --- | --- |
| predicted as ASD | TP | FP |
| not predicted as ASD | FN | TN |

**Table 4:**
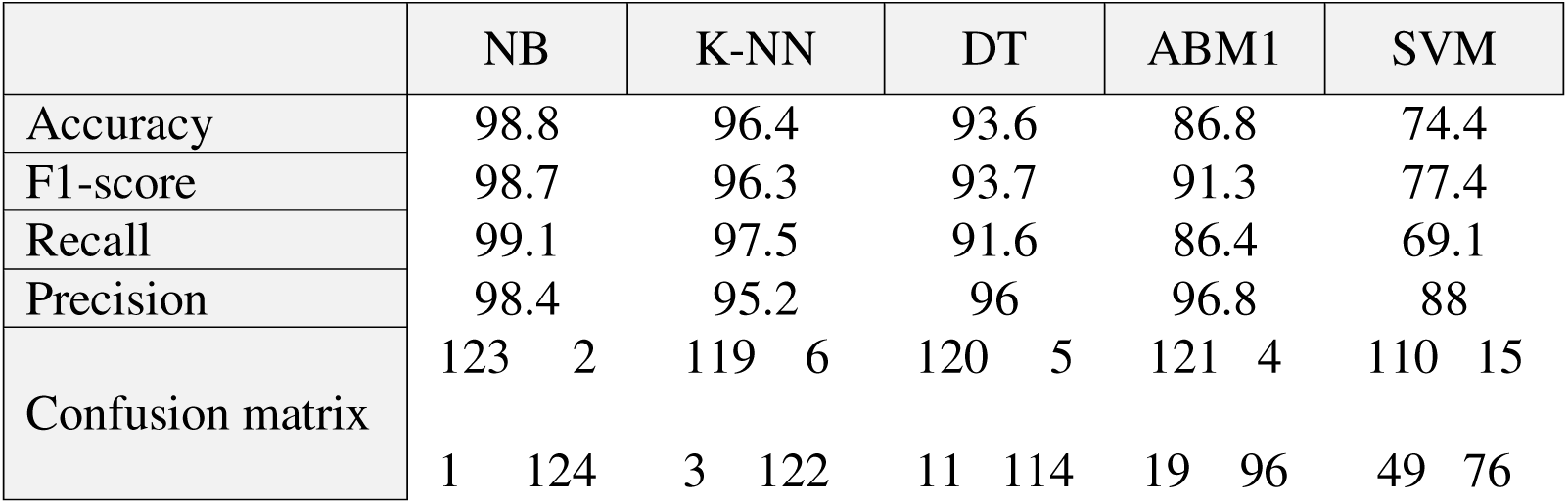
Comparison of evaluation metrics for tested algorithms based on ten features.

Precision, recall, and f-measure estimate interrater agreement from identified and predicted accuracy for qualitative variables were used to assess algorithm efficiency. Precision is a good assessment indicator when the proposed ML model must be validated using expected and actual results. It estimates predicted positive-to-actual positive ratio. This makes it dependent on TP and FP[20],[21].

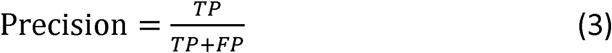

Recall, indicating the percentage of positives that were correctly classified, is another helpful assessment statistic to employ when trying to estimate how many positives may be reasonably expected. The values of TP and FN are used to quantify recall.

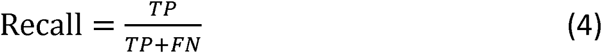

The F-Measure equilibrates accuracy and recall for a classifier. The F-Measure score is a value ranging from 0 to 1 that signifies the statistically relevant metrics of accuracy and recall.

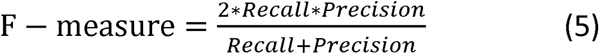

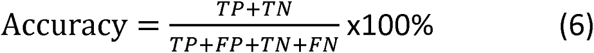

Were,

True positive (TP): The individual has ASD and it is predicted correctly.

True negative (TN): The individual does not have ASD and it is predicted correctly.

False positive (FP): The individual does not have ASD, but it is predicted incorrectly that the individual has ASD. This is known as Type 1 error.

False negative (FN): The individual has ASD, but it is predicted incorrectly that the individual does not have ASD. This is known as Type 2 error.

## 4. Results and Discussion

The evaluation details of the five implemented algorithms through 1250 cases for both ten features and after applying PCC to redact features to five distinct features (**sex, speech delay, jaundice, genetic disorder,** and **family_history**) introduced in Table (3 and 4) respectively.

The analysis of the two tables reveals a significant observation: reducing the features to half, based on the applied method, has demonstrated a substantial improvement across various metrics considered in the study. This indicates that feature selection plays a crucial role in enhancing the performance of predictive models by eliminating redundant or irrelevant features. By focusing on the most informative attributes, the model not only achieves higher accuracy and efficiency but also reduces computational complexity. This improvement underscores the importance of adopting robust down-sampling features techniques to optimize ML algorithms for better performance and reliability. Figure 2 illustrates the prediction accuracy of the five implemented algorithms for all ten features versus using the down-sampling features dataset.

**Fig 2:**
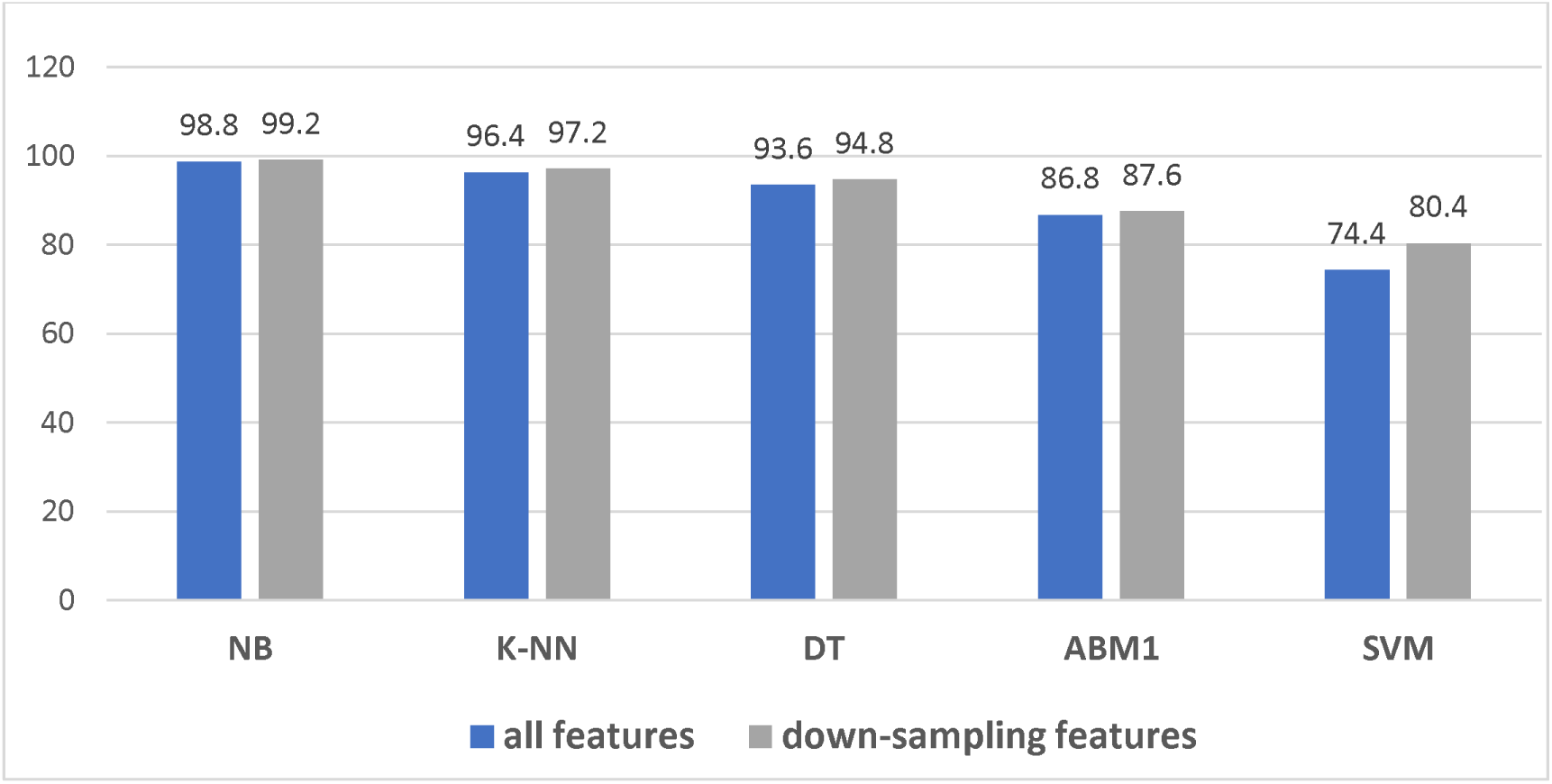
Prediction accuracy

**Fig 3:**
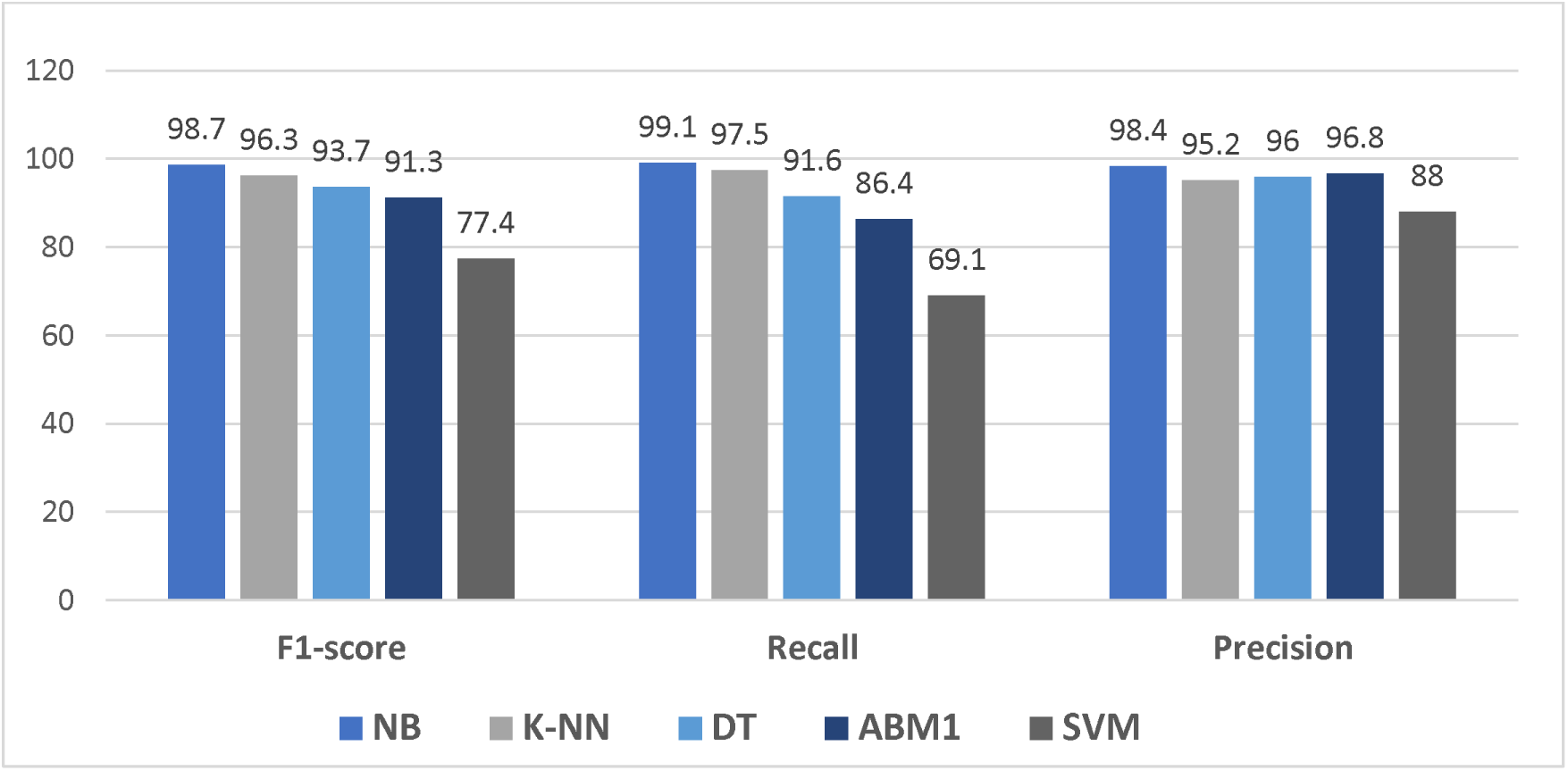
Performance metrics using ten features

In both scenarios, NB outperforms all other classifiers with an accuracy of 98.8%, 99.9%. the lowest accuracy of 74.4%, 80.4% by SVM algorithm.

F1-score, recall, precision and specificity of ten features as well as the down-sampling features are used to evaluate the results’ significance as demonstrated in Figure (3 and 4) respectively.

In the context of classification algorithms, the unit of time can refer to how measure the computational effort required to train and run these algorithms. Time for different ML algorithms vary widely depending on several factors such as: the size of the dataset, the complexity of the model, and the computational resources available. In this field, and to determine the efficiency of the implemented algorithms, the average time required to predict the outcome of each case of the data used for testing purposes and for each algorithm has been calculated. Inference or prediction time is the time it takes for a model to predict or infer the output for new data. The overall time required to procced ASD dataset for each of the implemented algorithms is illustrated in Figure 4.

**Fig 4:**
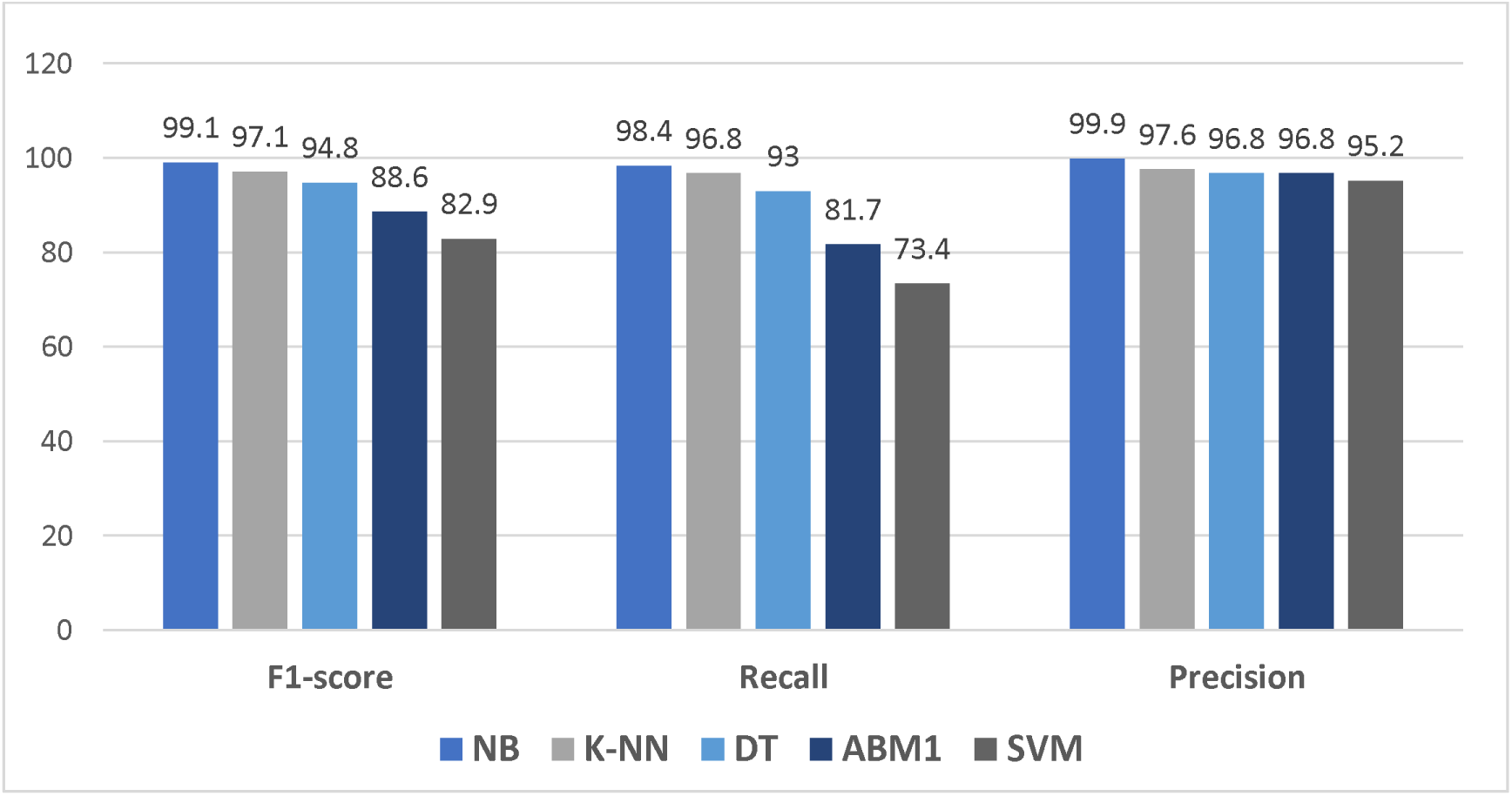
Performance metrics using down-sampling features

**Fig 4:**
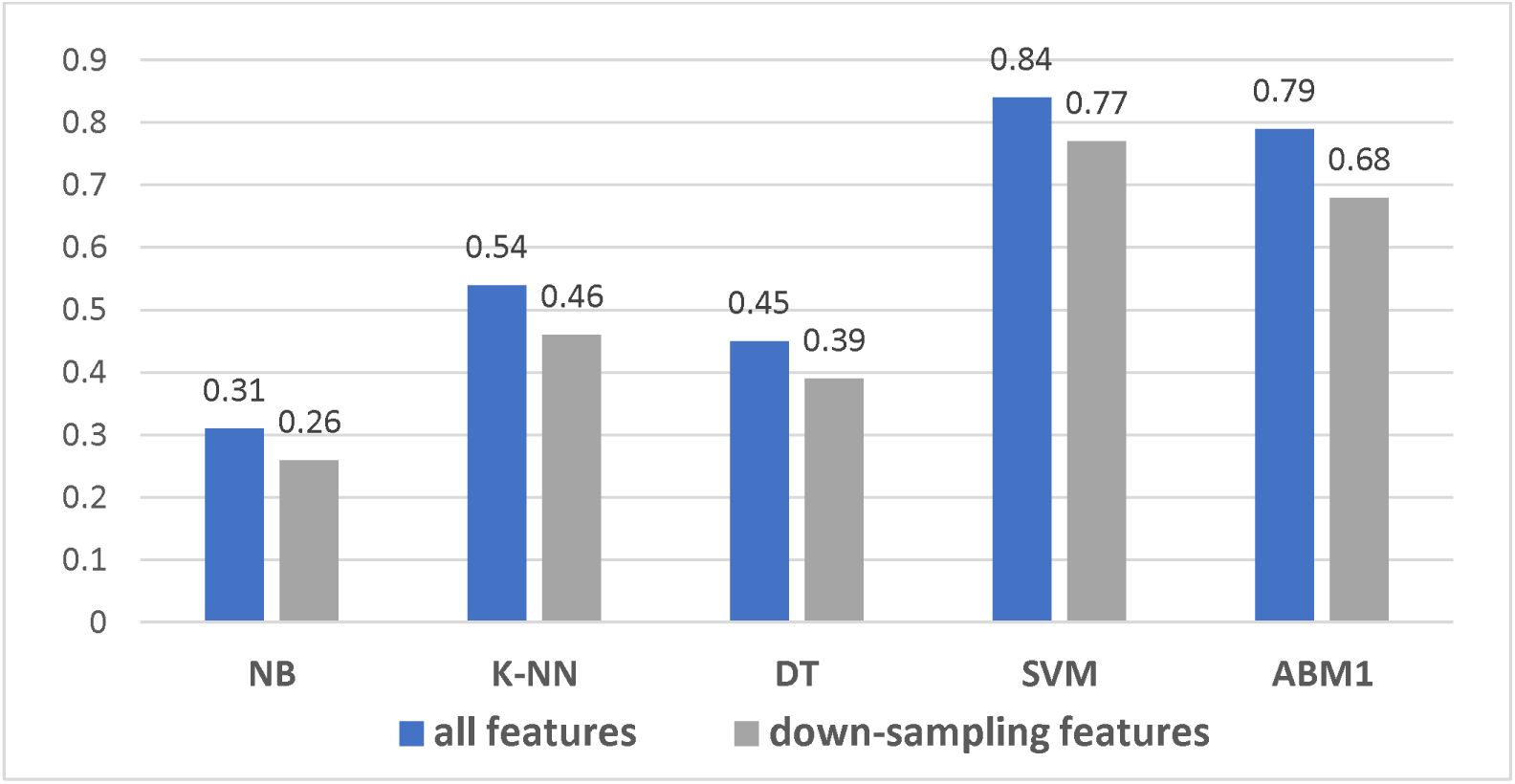
Time required to predict patient condition for the algorithms used

Clearly, the minimum prediction time of NB algorithm with 0.26 sec, meanwhile drastically increased to 0.46 sec with k-NN algorithm. However, the time has slightly decreased to 0.39 sec with DT algorithm, which is around half the recorded time of SVM as well as ABM1 algorithms.

The comparative results with other works show that better accuracy as well as prediction time can achieved by using proposed technique with down-sampling features. The number of reduced features also makes the test less sluggish. So, ML algorithms with feature reduction technique make the ASD prediction less time consuming along with better accuracy and the overall performance. A comparison of the proposed techniques with state of art works is presented in Table 6.

**Table 5:**
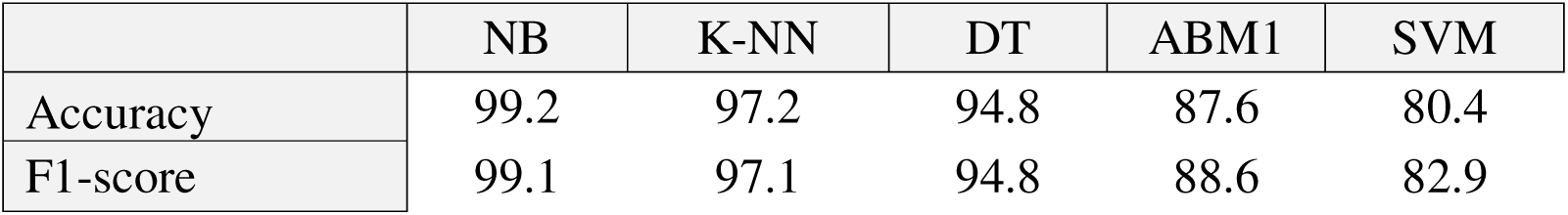
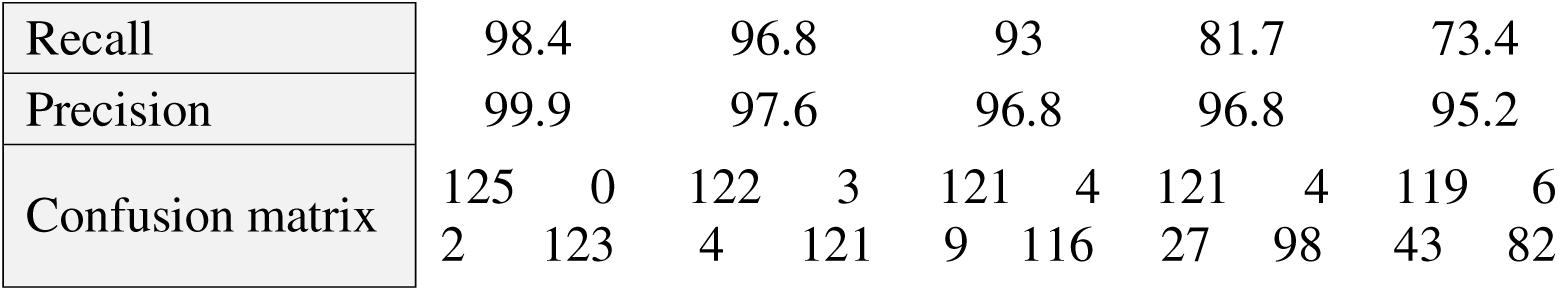
Comparison of evaluation metrics for tested algorithms based on five features.

|  | NB | K-NN | DT | ABM1 | SVM |
| --- | --- | --- | --- | --- | --- |
| Accuracy | 99.2 | 97.2 | 94.8 | 87.6 | 80.4 |
| F1-score | 99.1 | 97.1 | 94.8 | 88.6 | 82.9 |
| Recall | 98.4 | 96.8 | 93 | 81.7 | 73.4 |
| Precision | 99.9 | 97.6 | 96.8 | 96.8 | 95.2 |
| Confusion matrix | 125 0<br>2 123 | 122 3<br>4 121 | 121 4<br>9 116 | 121 4<br>27 98 | 119 6<br>43 82 |

**Table 6:** Comparison of the proposed models with previous works.

|  | Dataset | Method | Accuracy | Limitation | Strength |
| --- | --- | --- | --- | --- | --- |
| [7] | children records:292 | NB, SVM, LR, CNN, NN | NB:97.53%, SVM:96.30%, LR:96.88%, CNN:95%, NN:94.5% | does not predict the severity of ASD. In certain cases, the symptoms that are used to diagnose autism spectrum disorder (ASD) may not always be indicative of an actual instance of the disorder. | achieves high precision by utilizing five distinct ML-based categorization techniques. |
| [14] | children | LR, NB, SVM, | LR:93.1%<br>NB:94.7%<br>SVM:93.8% | As a consequence of the fact that the dataset was gathered from autism-based | forward feature selection was utilized, and five machine |
|  | records:1054<br>attributes:18 | KNN,<br>RF | KNN:90.5%<br>RF: 81.5% | collections for the most part, there was a rather considerable imbalance, especially in favor of the autism spectrum disorder class. | learning models were developed and assessed. |
| **Proposed method** | children records:1250<br>attributes: 5 | NB,<br>KNN,<br>DT<br>SVM,<br>ABM1 | NB:99.2%<br>KNN:97.2%<br>DT:94.8%<br>SVM:80.4%<br>ABM1:87.6% | ASD manifests differently in individuals, making it challenging to create a one-size-fits-all model. So, Rigorously validating models with diverse populations is crucial. | **Balanced** datasets of 1250 children have been processed for ASD prediction.<br>Implement **PCC** down-sampling features technique. |

In summary, the relationship between the number of features involved and prediction accuracy depends on the nature of the dataset and the quality of the features.

## 5. Conclusion

The significance of machine learning techniques might be utilized to forecast the appearance of the disease due to the fact that they have the capacity to accurately estimate the disease’s rate. In the present study, the efficiency of ML methods was tested for prediction using a 1250-record labeled balanced ASD dataset. In addition, the necessity of selecting the appropriate attributes in to get an accurate prediction was assessed for all of the algorithms that were implemented.Among several well-accepted, simple to construct, quick, and performed well algorithms, this work sought the most efficient ML approaches.

The experimental results revel that, NB algorithm yields superior performance compared to other four algorithms in terms of accuracy of 99.2% with minimum time whereas, SVM achieved the lowest accuracy of 80.4% with maximum time since when working with big datasets SVM implementations require extensive training time. Moreover, the result remarkably shows that by using Pearson correlation coefficient(PCC) feature reduction technique the overall performance of classifiers was enhanced since the number of attributes reduces to the half. ML algorithms processed with these selected features shows improvement in accuracy, sensitivity, specificity, and prediction time. The major objective of this work was to early and accurately predict of an important phenomenon in the future of childhood using MLmodels. In future, implement a hybrid model through a combination of ML algorithms with optimization techniques to produce an adaptive model for the diagnosis of ASD.

## Data Availability

All data produced are available online at Kaggle site

https://www.kaggle.com/datasets

## Acknowledgment

The author expresses sincere appreciation and gratitude to University of Baghdad. (www.uobaghdad.edu.iq) Baghdad-Iraq for its support.

## Author contributions

SWK conceptualized this article’s whole framework, data collection, analysis, and visualization. MAT contributed to supervising the work. Both authors participated in writing and revising the manuscript and approved the final manuscript.

## Ethical approval

The study was conducted with the approval of the Chairman of the scientific research ethics committee at the College of Science at the University of Baghdad, as well as the ethical committee’s review the implemented dataset and paper objectives. i.e. ethical approval was given. The study protocol was reviewed and approved by the scientific committee at the Faculty of Engineering - Department of Computer Engineering at the University of Baghdad.

## Informed consent

Informed consent was obtained from all subjects participating in the study.

